# Disability, Fatigue, and Mental Health in Acute versus Chronic Spinal Pain Patients in the Gaza Strip: A Comparative Cross-Sectional Study

**DOI:** 10.64898/2026.05.12.26353046

**Authors:** Marwa Salama, Ahmed A. Najim, Magdy Shabana, Reham Almukbel, Kinan Mokbel

## Abstract

**Background:** Spinal pain, including neck pain and low back pain (LBP), is a common musculoskeletal condition and major contributor to disability worldwide. Evidence comparing disability, fatigue and mental health across acute and chronic stages remains limited, particularly in conflict-affected and low-resource settings. This study assessed these outcomes among patients with acute and chronic neck pain and LBP in the Gaza Strip.

**Methods:** A comparative cross-sectional study was conducted among 410 adults attending outpatient physical therapy at Nasser Medical Complex, Khan Younis, Gaza Strip. Participants included 204 with neck pain and 206 with LBP, classified as acute neck pain (n=101), chronic neck pain (n=103), acute LBP (n=102) and chronic LBP (n=104). Disability, fatigue, psychological distress and sleep disturbance were assessed using the Neck Disability Index (NDI)/Oswestry Disability Index (ODI), Fatigue Severity Scale (FSS), Patient Health Questionnaire-4 (PHQ-4) and PROMIS Sleep Disturbance Short Form 8a. Independent t-tests, adjusted linear regression, correlation analyses, clinical-threshold analyses and binary logistic regression were performed.

**Results:** Chronic neck pain and chronic LBP were associated with significantly higher disability, fatigue and psychological distress than acute pain. Chronic neck pain patients had higher NDI, FSS and PHQ-4 scores than acute neck pain patients; chronic LBP patients had higher ODI, FSS and PHQ-4 scores than acute LBP patients (all *p*<0.001). Sleep disturbance did not differ significantly between groups. Female participants reported higher psychological distress in both pain groups, with higher fatigue in neck pain and higher disability in LBP. Adjusted analyses confirmed that chronic pain status remained associated with higher disability, fatigue and psychological distress. Fatigue was the most consistent factor independently associated with chronic pain status.

**Conclusions:** Chronic spinal pain was associated with greater disability, fatigue and psychological distress than acute spinal pain, while sleep disturbance was common across groups. These findings support early multidimensional assessment, including screening for fatigue and psychological distress. Longitudinal studies are needed to clarify whether these factors contribute to transition from acute to chronic spinal pain.

## Background

Low back pain (LBP) alone affected an estimated 619 million people worldwide in 2020 and is projected to affect 843 million people by 2050 [1]. Neck pain is also highly prevalent, with recent estimates showing more than 200 million prevalent cases globally and projections indicating that its population burden will remain substantial over time [2,3]. Together, LBP and neck pain are among the leading contributors to years lived with disability worldwide [4].

Spinal pain is commonly classified according to symptom duration, with acute pain usually defined as symptoms lasting up to 12 weeks and chronic pain as symptoms persisting beyond this period [5]. Although many patients recover during the acute phase, a substantial proportion develop persistent symptoms. In LBP, transition from acute to chronic pain has been reported in approximately one-third of patients in primary care settings [6]. Neck pain can also follow persistent trajectories over time, with some patients experiencing ongoing or recurrent symptoms rather than full recovery [7]. The mechanisms underlying this transition remain incompletely understood, although neurobiological and sensorimotor factors are increasingly recognised as relevant to persistent LBP [8].

Chronic spinal pain is rarely limited to pain alone. It is often accompanied by disability, fatigue, psychological distress and sleep disturbance. Disability, as framed within the International Classification of Functioning, Disability and Health, includes impairments, activity limitations and participation restrictions [9]. In spinal pain, disability is clinically important because it reflects the extent to which pain affects daily functioning, work, self-care and participation. Previous studies have identified disability as an important feature of chronic neck pain and have shown that patients with chronic neck and LBP experience greater functional limitation and emotional burden [10,11].

Fatigue is another important but often under-assessed feature of chronic spinal pain. In chronic LBP, fatigue has been linked with pain intensity, depressive symptoms and disability [12]. Evidence from chronic musculoskeletal pain populations also suggests a temporal relationship between pain and fatigue, although the direction of this relationship is likely complex and bidirectional [13]. This makes fatigue a clinically relevant outcome when comparing acute and chronic spinal pain groups.

Psychological distress is also central to the chronic spinal pain experience. Depression and anxiety are closely associated with persistent pain, and longitudinal evidence suggests that chronic LBP is linked with increased risk of depression and anxiety symptoms [14]. A systematic review of prognostic factors for LBP chronicity also identified psychological factors, including depression and anxiety, as important contributors to persistent pain [15]. More broadly, clinically significant depressive and anxiety symptoms are common among adults with chronic pain [16]. These findings support the need to assess mental health alongside physical disability in patients with spinal pain.

Sleep disturbance is another relevant domain. Poor sleep may be both a consequence of pain and a factor that contributes to poorer pain outcomes. Prospective evidence suggests that sleep can act as a prognostic factor in LBP outcomes [17], and systematic-review evidence supports a bidirectional relationship between sleep problems and chronic musculoskeletal pain [18]. However, the relationship between sleep and spinal pain may be influenced by wider contextual factors, especially in settings affected by conflict, insecurity and displacement.

Sex differences should also be considered when studying spinal pain outcomes. Women have been reported to experience a higher prevalence of chronic neck pain and chronic LBP than men [19]. Broader literature on chronic musculoskeletal conditions also highlights sex-related differences in pain prevalence, pain experience and associated outcomes [20]. Gender-related issues in LBP management may reflect a combination of biological, psychosocial and healthcare-access factors [21]. Recent evidence further suggests that sex may influence the relationship between pain characteristics, disability and quality of life in people with chronic spinal pain [22].

Most previous studies have examined neck pain and LBP separately, or have focused on a single outcome such as disability, psychological distress or sleep. Fewer studies have assessed disability, fatigue and mental health outcomes simultaneously across acute and chronic stages of both neck pain and LBP. Evidence from conflict-affected and low-resource settings is especially limited, despite evidence that displacement, trauma and migration-related stressors are associated with chronic pain, psychological distress and functional impairment [23–25].

The Gaza Strip provides an important context for this research. Since October 2023, the ongoing conflict has severely disrupted healthcare access and intensified population-level psychological distress. A United Nations humanitarian update citing the World Health Organization reported that the number of people in Gaza requiring mental health and psychosocial support had increased from approximately 485,000 before October 2023 to more than one million by 2025 [26]. In this context, patients with spinal pain may experience a compounded burden shaped by physical symptoms, psychological stress, displacement and limited access to rehabilitation.

This study therefore aimed to compare disability, fatigue and mental health outcomes, including psychological distress and sleep disturbance, among patients with acute and chronic neck pain and LBP attending a major tertiary hospital in the Gaza Strip. The study also examined sex differences in these outcomes and explored factors associated with chronic spinal pain status.

## Methods

### Study design and setting

A comparative cross-sectional study was conducted at the outpatient physical therapy department of Nasser Medical Complex, Khan Younis, Gaza Strip, Palestine. This design was selected because the study aimed to compare disability, fatigue and mental health outcomes between patients with acute and chronic spinal pain at a single time point. The design was therefore appropriate for identifying group differences and associations, but not for establishing temporal or causal relationships.

Nasser Medical Complex is one of the main governmental hospitals in Gaza and provides outpatient physiotherapy services for patients with musculoskeletal conditions, including neck pain and LBP. Data were collected between mid-August 2025 and December 2025.

### Participants

The study population comprised adults aged 18 years or older who attended the outpatient physical therapy department with neck pain or LBP. Participants were classified by pain location and duration into four groups: acute neck pain, chronic neck pain, acute LBP and chronic LBP. Pain duration was recorded using a 12-week cut-off. Acute spinal pain was defined as symptoms lasting 12 weeks or less, while chronic spinal pain was defined as symptoms persisting for more than 12 weeks [5].

Patients were excluded if they had a current or previous autoimmune or systemic inflammatory disease, current or previous malignancy, spinal surgery within the preceding six months, previous spinal trauma, congenital or acquired spinal defect, pregnancy, comorbid neurological disorder, severe cognitive impairment, or current or previous dependence on systemic corticosteroids.

### Sampling and sample size

A non-probability consecutive sampling method was used. Patients presenting to the outpatient physical therapy department with neck pain or LBP were screened for eligibility, and eligible patients were invited to participate during the data collection period. The target sample size was calculated using OpenEpi version 3 for estimating a frequency in a population. The calculation assumed a population size of 1,000,000, an expected outcome frequency of 32%, confidence limits of 5%, a design effect of 1 and a confidence level of 97%, giving a required sample size of 410 participants. The expected frequency was informed by previously reported transition from acute to chronic LBP [6]. The final analysed sample included 410 participants: 204 with neck pain and 206 with LBP. Participants were classified into four pain-location and pain-duration groups: acute neck pain (n=101), chronic neck pain (n=103), acute LBP (n=102) and chronic LBP (n=104). A pilot study involving 41 participants, equivalent to 10% of the final sample size, was conducted to assess the clarity and applicability of the questionnaire and study procedures. Pilot participants were not included in the final analysis.

### Outcome measures

Disability was assessed using adapted region-specific instruments. For participants with neck pain, the Neck Disability Index (NDI) was used. The NDI is a ten-item self-report measure of neck-related disability, with higher scores indicating greater disability [27]. For participants with LBP, the Oswestry Disability Index (ODI) was used. The ODI is a widely used condition-specific measure of LBP-related functional limitation, with higher scores indicating greater disability [28]. In the administered versions, the NDI driving item and the ODI travelling item were excluded from the total score because these activities were not consistently applicable during the conflict context in Gaza, when access to vehicles and ordinary travel was severely restricted. Both measures were therefore analysed as nine-item raw total scores, with possible scores ranging from 0 to 45. Higher scores indicated greater disability.

Fatigue was assessed using the Fatigue Severity Scale (FSS), a nine-item measure evaluating the impact of fatigue on daily functioning. Each item is scored from 1 to 7, giving a total score range of 9 to 63. Higher scores indicate greater fatigue severity. A mean item score of 4 or above, equivalent to a total score of 36 or above, was considered clinically significant fatigue [29].

Psychological distress was assessed using the Patient Health Questionnaire-4 (PHQ-4), a brief four-item screening tool that includes two items assessing anxiety symptoms and two items assessing depressive symptoms. Total scores range from 0 to 12, with higher scores indicating greater psychological distress. Brief depression and anxiety screening measures have demonstrated utility in chronic pain and persistent musculoskeletal pain populations [30,31], and the assessment of psychological outcomes is consistent with recommended approaches for chronic musculoskeletal pain research [32].

Sleep disturbance was assessed using the PROMIS Sleep Disturbance Short Form 8a. The measure includes eight items assessing perceived sleep quality and sleep-related difficulties. In this study, PROMIS sleep disturbance was analysed using the raw summed score, with higher scores indicating greater sleep disturbance.

Internal consistency was acceptable for all instruments in the study sample. Cronbach’s alpha values were 0.818 for NDI, 0.771 for ODI, 0.844 for FSS, 0.777 for PHQ-4 and 0.775 for PROMIS Sleep Disturbance. All questionnaires and study instruments are provided in the Supplementary Materials (Additional file 1).

### Data collection

Eligible participants were approached in the outpatient physical therapy department. After providing written informed consent, participants completed a structured questionnaire through researcher-administered interview. Separate questionnaires were used for neck pain and LBP participants, with each questionnaire including sociodemographic information and the relevant outcome measures.

The sociodemographic and clinical variables collected included sex, age group, employment status, pain duration and previous physiotherapy for the current complaint. Pain duration was used to classify participants as having acute or chronic pain.

### Statistical analysis

Data were analysed using Stata/SE version 19 (StataCorp, College Station, TX, USA). Missing questionnaire responses were minimal. Analyses were conducted using available data for each outcome, with valid denominators reported in the tables. Descriptive statistics were used to summarise participant characteristics and outcome measures. Categorical variables were reported as frequencies and percentages, while continuous variables were reported as mean ± standard deviation.

Independent samples t-tests were used to compare disability, fatigue, psychological distress and sleep disturbance between acute and chronic pain groups. Separate comparisons were conducted for neck pain and LBP because disability was measured using different region-specific instruments. Independent samples t-tests were also used to examine sex differences in outcome measures within the neck pain and LBP groups.

Pearson correlation coefficients were calculated to examine associations between sleep disturbance, fatigue, disability and psychological distress within each pain-location group. Binary logistic regression analyses were then conducted separately for neck pain and LBP to identify factors independently associated with chronic pain status. In these models, pain duration was the dependent variable and was coded as acute pain = 0 and chronic pain = 1. For the neck pain model, PROMIS, FSS, PHQ-4 and NDI were entered as independent variables. For the LBP model, PROMIS, FSS, PHQ-4 and ODI were entered as independent variables.

Additional analyses were conducted to assess the robustness and clinical interpretability of the findings. Adjusted linear regression models were used to examine whether acute/chronic differences in outcome scores remained after adjustment for sex, age group, employment status and previous physiotherapy. Standardised mean differences were calculated using Cohen’s d to estimate the magnitude of between-group differences. Additional categorical analyses were conducted using established thresholds for clinically significant fatigue and PHQ-4 psychological distress categories. Because questionnaire scores may not be normally distributed, Mann-Whitney U tests and Spearman rank correlations were conducted as sensitivity analyses. Exploratory sex-by-pain-status interaction models were also conducted to examine whether sex modified the association between pain status and each outcome.

Because the study was cross-sectional, regression findings were interpreted as associations with chronic pain status rather than evidence of temporal prediction or causation. Statistical significance was set at *p*≤0.05. Because the primary analyses were prespecified and clinically defined, *p*-values were not routinely adjusted for multiple testing. Findings from exploratory interaction and sensitivity analyses were interpreted cautiously, with attention to the number of comparisons.

### Use of large language models

ChatGPT was used to support manuscript language editing. The authors verified the final manuscript content and take full responsibility for the work. No large language model generated primary data or was listed as an author.

### Ethical considerations

Ethical approval was granted by the Helsinki Committee for Ethical Approval, Palestinian Health Research Council, Gaza Strip, Palestine (approval number PHRC/HC/1240/25; 04 August 2025). Administrative approval was obtained from the Ministry of Health, State of Palestine, Human Resources Development Directorate (correspondence number 25114150; 13 August 2025). Permission to conduct the research was also obtained from Al-Azhar University on 31 July 2025. Written informed consent was obtained from all participants before participation. All personal information was treated confidentially and was accessible only to the research team.

## Results

### Sample characteristics

The study included 410 participants attending the outpatient physical therapy department with either neck pain or LBP. Of these, 204 participants were in the neck pain group and 206 were in the LBP group. Participants were then classified according to pain duration using the 12-week cut-off, with acute pain defined as symptoms lasting ≤12 weeks and chronic pain defined as symptoms lasting >12 weeks. This resulted in four groups: acute neck pain (n=101), chronic neck pain (n=103), acute LBP (n=102) and chronic LBP (n=104) (Figure 1). Overall, 203 participants had acute spinal pain and 207 had chronic spinal pain. Missing outcome data were minimal, and valid denominators are reported in the relevant tables.

**Figure 1.**
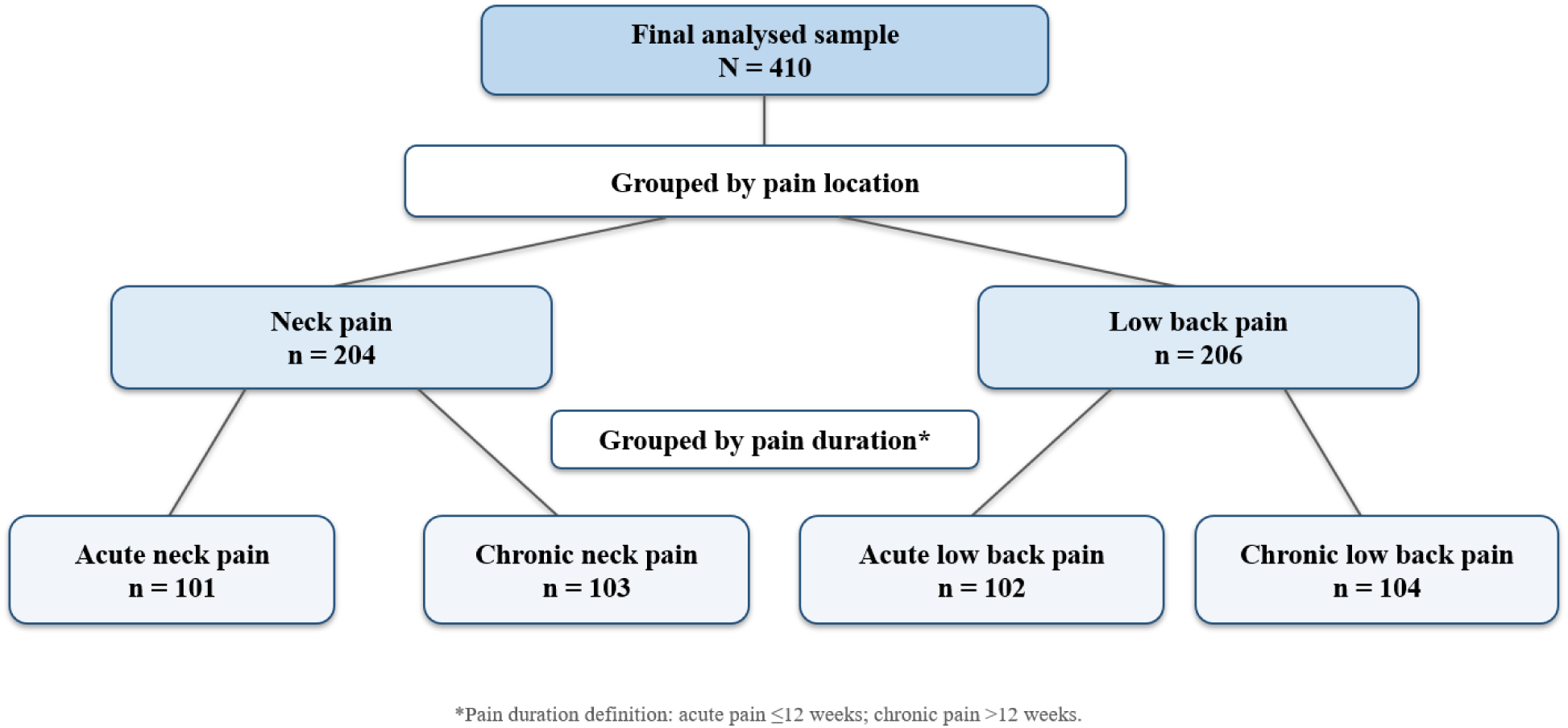
Flow chart of participant grouping by pain location and pain duration. Participants were first grouped by pain location into neck pain and low back pain groups, and then by pain duration into acute pain (≤12 weeks) and chronic pain (>12 weeks).

Females represented the majority of the sample (n=265, 64.6%), while 145 participants were male (35.4%). The most common age group was 21-30 years (n=137, 33.4%), followed by 41-50 years (n=123, 30.0%) and 31-40 years (n=117, 28.5%). More than half of participants were unemployed (n=227, 55.4%), and most had not received previous physiotherapy for their current complaint (n=298, 72.7%) (Supplementary Table S1; Additional file 1).

### Disability in acute and chronic spinal pain

Patients with chronic spinal pain reported significantly higher disability scores than those with acute spinal pain (Table 1). In the neck pain group, patients with chronic neck pain had higher NDI scores than those with acute neck pain (21.59 ± 7.78 vs 11.98 ± 5.46; t=10.202, *p*<0.001). A similar pattern was observed in the LBP group, where patients with chronic LBP had higher ODI scores than those with acute LBP (17.29 ± 7.36 vs 12.09 ± 5.64; t=5.684, *p*<0.001).

**Table 1.** Mean differences in disability scores between acute and chronic spinal pain groups.

| Outcome measure | Acute pain Mean ± SD | Chronic pain Mean ± SD | t | p-value | Cohen's d |
| --- | --- | --- | --- | --- | --- |
| NDI, neck pain | 11.98 ± 5.46 (n=101) | 21.59 ± 7.78 (n=103) | -10.202 | <0.001 | 1.43 |
| ODI, low back pain | 12.09 ± 5.64 (n=102) | 17.29 ± 7.36 (n=104) | -5.684 | <0.001 | 0.79 |

Note: Values are mean ± SD. NDI and ODI were analysed as adapted nine-item raw total scores ranging from 0 to 45.
NDI, Neck Disability Index; ODI, Oswestry Disability Index; SD, standard deviation.

These findings indicate that chronic pain status was associated with greater functional limitation in both neck pain and LBP groups.

### Fatigue in acute and chronic spinal pain

Fatigue scores were significantly higher among participants with chronic pain than those with acute pain in both pain-location groups (Table 2). In the neck pain group, the chronic pain subgroup had a higher mean FSS score than the acute pain subgroup (42.10 ± 8.09 vs 27.63 ± 13.27; t=9.385, *p*<0.001). Similarly, in the LBP group, patients with chronic pain had higher FSS scores than those with acute pain (41.89 ± 10.70 vs 29.13 ± 14.14; t=7.318, *p*<0.001).

**Table 2.** Mean differences in fatigue severity between acute and chronic spinal pain groups.

| Outcome measure | Acute pain Mean ± SD | Chronic pain Mean ± SD | t | p-value | Cohen's d |
| --- | --- | --- | --- | --- | --- |
| FSS, neck pain | 27.63 ± 13.27 (n=101) | 42.10 ± 8.09 (n=102) | -9.385 | <0.001 | 1.32 |
| FSS, low back pain | 29.13 ± 14.14 (n=102) | 41.89 ± 10.70 (n=104) | -7.318 | <0.001 | 1.02 |

Note: Values are mean ± SD. FSS, Fatigue Severity Scale; SD, standard deviation.

The mean FSS scores in both chronic pain groups exceeded the clinically significant fatigue threshold, equivalent to a mean item score of 4 or above.

### Psychological distress and sleep disturbance

Psychological distress was significantly higher in chronic pain groups than acute pain groups (Table 3). Among patients with neck pain, PHQ-4 scores were higher in the chronic group than in the acute group (6.94 ± 2.79 vs 3.64 ± 3.17; t=7.880, *p*<0.001). The same pattern was observed among patients with LBP, where the chronic group reported higher PHQ-4 scores than the acute group (6.31 ± 3.64 vs 4.00 ± 3.31; t=4.761, *p*<0.001).

**Table 3.** Mean differences in psychological distress and sleep disturbance between acute and chronic spinal pain groups.

| Measure | Group | Acute pain Mean ± SD | Chronic pain Mean ± SD | t | p-value | Cohen's d |
| --- | --- | --- | --- | --- | --- | --- |
| PHQ-4 | Neck pain | 3.64 ± 3.17 (n=100) | 6.94 ± 2.79 (n=103) | -7.880 | <0.001 | 1.11 |
| PHQ-4 | Low back pain | 4.00 ± 3.31 (n=102) | 6.31 ± 3.64 (n=104) | -4.761 | <0.001 | 0.66 |
| PROMIS Sleep Disturbance | Neck pain | 25.23 ± 4.87 (n=100) | 25.14 ± 5.73 (n=102) | 0.124 | 0.902 | -0.02 |
| PROMIS Sleep Disturbance | Low back pain | 24.52 ± 4.84 (n=102) | 25.91 ± 6.95 (n=104) | -1.667 | 0.097 | 0.23 |

Note: Values are mean ± SD. PHQ-4, Patient Health Questionnaire-4; PROMIS, PROMIS Sleep Disturbance Short Form 8a raw summed score; SD, standard deviation.

In contrast, PROMIS sleep disturbance scores did not differ significantly between acute and chronic pain groups. In the neck pain group, PROMIS scores were almost identical between acute and chronic pain patients (25.23 ± 4.87 vs 25.14 ± 5.73; t=0.124, *p*=0.902). In the LBP group, PROMIS scores were slightly higher in the chronic group, but the difference was not statistically significant (25.91 ± 6.95 vs 24.52 ± 4.84; t=1.667, *p*=0.097).

These findings suggest that psychological distress differed clearly by pain duration, whereas sleep disturbance was present across groups regardless of acute or chronic pain status.

### Sex differences in disability, fatigue and mental health outcomes

Sex-based comparisons are presented in Supplementary Tables S2 and S3 (Additional file 1). In the neck pain group, females reported significantly higher fatigue than males (37.17 ± 12.44 vs 29.98 ± 13.36; t=3.734, *p*<0.001). Females also reported higher psychological distress, as reflected by PHQ-4 scores (5.76 ± 3.39 vs 4.36 ± 3.27; t=2.757, *p*=0.006). There were no significant sex differences in NDI or PROMIS scores among patients with neck pain.

In the LBP group, females had significantly higher ODI scores than males (15.66 ± 7.14 vs 13.25 ± 6.68; t=2.433, *p*=0.016). Females also reported higher psychological distress than males (PHQ-4: 5.62 ± 3.66 vs 4.46 ± 3.56; t=2.260, *p*=0.025). No significant sex differences were observed for FSS or PROMIS scores in the LBP group.

Overall, females appeared to report higher psychological distress in both pain-location groups, with additional unadjusted differences in fatigue among neck pain patients and disability among LBP patients.

### Factors associated with chronic pain status

Binary logistic regression analyses were conducted separately for neck pain and LBP groups (Tables 4 and 5). In the neck pain model, higher FSS scores were independently associated with chronic neck pain status (OR=1.061, 95% CI 1.022-1.102; *p*=0.002). Higher NDI scores were also independently associated with chronic neck pain status (OR=1.148, 95% CI 1.079-1.220; *p*<0.001). PROMIS sleep disturbance scores showed a negative association with chronic neck pain status (OR=0.907, 95% CI 0.846-0.974; *p*=0.007), while PHQ-4 was not independently associated with chronic neck pain status after adjustment for the other variables in the model (*p*=0.112).

**Table 4.** Binary logistic regression examining factors associated with chronic neck pain status.

| Variable | B | SE | Wald | p-value | OR (95% CI) |
| --- | --- | --- | --- | --- | --- |
| PROMIS | -0.097 | 0.036 | 7.241 | 0.007 | 0.907 (0.846-0.974) |
| FSS | 0.060 | 0.019 | 9.666 | 0.002 | 1.061 (1.022-1.102) |
| PHQ-4 | 0.111 | 0.070 | 2.530 | 0.112 | 1.118 (0.974-1.282) |
| NDI | 0.138 | 0.031 | 19.348 | <0.001 | 1.148 (1.079-1.220) |

Note: Model n=201 complete cases. OR, odds ratio; CI, confidence interval; SE, standard error; PROMIS, PROMIS Sleep Disturbance Short Form 8a raw summed score; FSS, Fatigue Severity Scale; PHQ-4, Patient Health Questionnaire-4; NDI, Neck Disability Index. NDI was analysed as an adapted nine-item raw total score ranging from 0 to 45. B represents the unstandardised logistic regression coefficient on the log-odds scale.

**Table 5.**
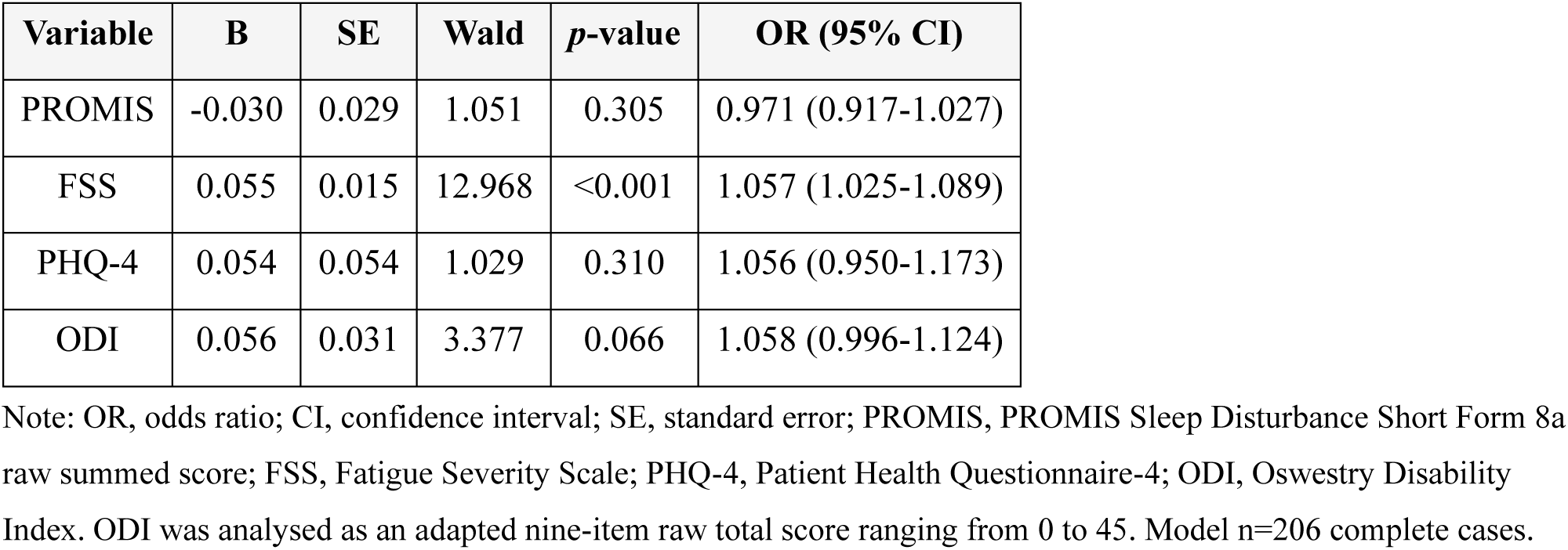
Binary logistic regression examining factors associated with chronic low back pain status.

| Variable | B | SE | Wald | p-value | OR (95% CI) |
| --- | --- | --- | --- | --- | --- |
| PROMIS | -0.030 | 0.029 | 1.051 | 0.305 | 0.971 (0.917-1.027) |
| FSS | 0.055 | 0.015 | 12.968 | <0.001 | 1.057 (1.025-1.089) |
| PHQ-4 | 0.054 | 0.054 | 1.029 | 0.310 | 1.056 (0.950-1.173) |
| ODI | 0.056 | 0.031 | 3.377 | 0.066 | 1.058 (0.996-1.124) |

Note: OR, odds ratio; CI, confidence interval; SE, standard error; PROMIS, PROMIS Sleep Disturbance Short Form 8a raw summed score; FSS, Fatigue Severity Scale; PHQ-4, Patient Health Questionnaire-4; ODI, Oswestry Disability Index. ODI was analysed as an adapted nine-item raw total score ranging from 0 to 45. Model n=206 complete cases.

In the LBP model, FSS was the only variable independently associated with chronic LBP status (OR=1.057, 95% CI 1.025-1.089; *p*<0.001). ODI approached statistical significance but did not reach the *p*≤0.05 threshold (OR=1.058, 95% CI 0.996-1.124; *p*=0.066). PROMIS and PHQ-4 were not independently associated with chronic LBP status in this model.

Given the cross-sectional design, these findings should be interpreted as associations with chronic pain status rather than evidence of temporal prediction or causation.

### Correlations among outcome variables

Correlation analyses showed significant positive associations among the main outcome variables in both pain-location groups (Supplementary Tables S4 and S5; Additional file 1). In the neck pain group, fatigue was strongly correlated with psychological distress (r=0.657, *p*<0.001) and disability (r=0.577, *p*<0.001). Disability was also strongly correlated with psychological distress (r=0.568, *p*<0.001). PROMIS sleep disturbance showed weaker but statistically significant correlations with fatigue, disability and psychological distress.

A similar pattern was observed in the LBP group. Fatigue was strongly correlated with disability (r=0.604, *p*<0.001) and psychological distress (r=0.553, *p*<0.001). Disability was moderately correlated with psychological distress (r=0.481, *p*<0.001). PROMIS sleep disturbance was also significantly correlated with fatigue, disability and psychological distress.

Together, these findings indicate that fatigue, disability, psychological distress and sleep disturbance are interrelated in both neck pain and LBP patients, although sleep disturbance did not differ significantly between acute and chronic groups.

### Additional and sensitivity analyses

Additional adjusted linear regression models showed that chronic pain status remained significantly associated with higher disability, fatigue and psychological distress in both neck pain and LBP groups after adjustment for sex, age group, employment status and previous physiotherapy (Supplementary Table S6; Additional file 1). Chronic pain status was not significantly associated with PROMIS sleep disturbance scores after adjustment in either pain-location group.

Clinical-threshold analyses showed that clinically significant fatigue was more common in chronic than acute pain groups for both neck pain and LBP. Moderate or severe psychological distress was also more frequent in chronic pain groups (Supplementary Table S7; Additional file 1).

Sensitivity analyses using Mann-Whitney U tests supported the primary findings for disability, fatigue and psychological distress (Supplementary Table S8; Additional file 1). PROMIS findings were not fully consistent across primary and sensitivity analyses and were therefore interpreted cautiously. Spearman correlation analyses broadly supported the Pearson correlation findings, particularly the strong associations between fatigue, disability and psychological distress (Supplementary Table S9; Additional file 1).

Exploratory sex-by-pain-status interaction analyses suggested that sex may modify the association between pain status and both fatigue and psychological distress in the neck pain group, but not in the LBP group (Supplementary Table S10; Additional file 1). These interaction findings should be interpreted cautiously because the study was not primarily powered for interaction testing.

## Discussion

### Principal findings

This comparative cross-sectional study found that chronic neck pain and chronic LBP were associated with significantly higher disability, fatigue and psychological distress than acute pain. Sleep disturbance showed a different pattern: it did not differ significantly between acute and chronic groups, although it was correlated with fatigue, disability and psychological distress in both pain-location groups. In the location-specific regression models, fatigue was the most consistent factor independently associated with chronic pain status. Neck disability was also independently associated with chronic neck pain status, while LBP disability approached but did not reach statistical significance.

Additional adjusted analyses strengthened the main findings. Chronic pain status remained associated with higher disability, fatigue and psychological distress after adjustment for basic participant characteristics including sex, age group, employment status and previous physiotherapy. The clinical-threshold analyses also showed that chronic pain groups had a greater burden of clinically significant fatigue and moderate/severe psychological distress. These findings suggest that the observed differences are not only statistically significant but also clinically meaningful.

These findings support a multidimensional interpretation of chronic spinal pain. Chronicity was not reflected only in higher disability scores, but also in greater fatigue and psychological burden. This is clinically important because it suggests that patients with chronic spinal pain may require assessment beyond pain location and pain duration alone. However, because the study was cross-sectional, the findings should be interpreted as associations with chronic pain status rather than evidence of causation or temporal prediction.

### Disability and chronic spinal pain

Disability was significantly higher in chronic neck pain and chronic LBP than in the corresponding acute groups. This indicates that persistent spinal pain is associated with greater functional limitation across both spinal regions. Similar findings have been reported in chronic back pain populations, where higher pain intensity and disability were associated with depressive symptoms and poorer functioning [33]. The present study adds to this evidence by showing that higher disability is evident when acute and chronic groups are compared directly within the same clinical setting.

In the adjusted models, disability remained independently associated with chronic pain status in the neck pain group, while ODI approached but did not reach statistical significance in the LBP group. This suggests that disability was clearly higher in chronic pain groups, but its independent association with chronicity differed by spinal region after adjustment for fatigue, psychological distress and sleep disturbance.

Disability in spinal pain is unlikely to be explained by pain duration alone. It is more likely to reflect the interaction between persistent pain, reduced activity, emotional distress and avoidance of movement. Previous research has shown that catastrophising, fear of movement, anxiety and depression are associated with persistent severe LBP and disability [34]. A systematic review of mediation studies also suggested that psychological distress and fear may partly explain the pathway between pain and disability in musculoskeletal pain [35]. This supports the interpretation that disability in chronic spinal pain is a biopsychosocial outcome rather than a purely physical consequence.

The findings are also consistent with research in non-specific LBP showing that disability, fatigue and psychological factors vary according to the degree of chronification [36]. In the current study, a similar multidomain pattern was visible across both neck pain and LBP. Clinically, this suggests that disability screening should be included early in the assessment of spinal pain, especially when symptoms persist beyond the acute stage.

### Fatigue as a central feature of chronic spinal pain

Fatigue was the most consistent finding across the primary and additional analyses. It differed significantly between acute and chronic groups, remained significant in the adjusted linear models, and was the only factor independently associated with chronic pain status in both location-specific logistic regression models. Patients with chronic neck pain and chronic LBP had substantially higher FSS scores than patients with acute pain. Both chronic pain groups also had mean fatigue scores above the clinically significant threshold, suggesting that fatigue is not a minor accompanying symptom but a central feature of the chronic spinal pain burden.

This finding is supported by previous evidence showing that patients with chronic LBP and chronic neck pain report elevated fatigue compared with non-pain comparison groups [37]. In neck pain specifically, fatigue may also affect neuromuscular control. Hsu et al. reported that fatigue altered neck muscle control and worsened postural stability during arm movement perturbations in patients with chronic neck pain [38]. This provides a plausible mechanism through which fatigue may contribute to functional limitation in chronic neck pain.

In the present study, fatigue was strongly correlated with disability and psychological distress in both pain-location groups. This is important clinically. Fatigue may reduce engagement with rehabilitation, limit activity tolerance and increase perceived disability. It may also interact with psychological distress, creating a cycle in which persistent pain, low energy, reduced activity and emotional symptoms reinforce one another. Routine fatigue screening may therefore help identify patients who need closer follow-up, pacing advice and broader rehabilitation support.

### Psychological distress and chronic spinal pain

Psychological distress was significantly higher in chronic pain groups than in acute pain groups. Patients with chronic neck pain and chronic LBP both reported higher PHQ-4 scores than those with acute pain. This supports the view that chronic spinal pain is associated with a greater emotional burden.

However, PHQ-4 did not remain independently associated with chronic pain status in the adjusted location-specific regression models. Psychological distress should therefore be interpreted as an important marker of clinical burden rather than an independent explanatory factor in these models.

The neck pain findings are consistent with previous work showing that psychological states are associated with pain and disability in chronic neck pain patients [39]. High levels of anxiety and depression have also been reported among patients with chronic neck pain, highlighting the importance of psychological assessment in this population [40]. In the current study, psychological distress was strongly correlated with disability and fatigue, suggesting that emotional symptoms are closely connected to the broader clinical burden of chronic spinal pain.

However, psychological distress should not be interpreted simply as a consequence of spinal pain. In Gaza, ongoing conflict, displacement, insecurity, bereavement and restricted access to care may contribute to psychological distress across the whole population. This may partly explain why psychological distress was also present in acute pain groups. The safer interpretation is that chronic spinal pain status was associated with higher psychological distress within an already highly stressed context.

### Sleep disturbance across acute and chronic groups

Sleep disturbance did not differ significantly between acute and chronic pain groups. This contrasts with previous evidence suggesting an association between sleep and chronic spinal pain [41]. Systematic review and meta-analysis evidence also supports a bidirectional relationship between chronic musculoskeletal pain and sleep-related problems [42]. In LBP specifically, cohort evidence has suggested bidirectional associations between chronic LBP and sleep quality over time [43].

The absence of a significant difference in the present study does not mean that sleep is unimportant. PROMIS sleep disturbance scores were significantly correlated with fatigue, disability and psychological distress in both neck pain and LBP groups. The negative association between PROMIS score and chronic neck pain status in the adjusted model should be interpreted cautiously, particularly because sleep disturbance did not differ significantly between acute and chronic neck pain groups in the unadjusted comparison. This finding may reflect adjustment effects or shared contextual sleep disruption across the sample rather than a clear protective effect of poorer sleep. In Gaza, sleep may be disrupted by insecurity, displacement, overcrowding, environmental noise and psychological stress. These background stressors may mask differences between acute and chronic pain groups.

This finding matters because sleep disturbance may function differently in conflict-affected settings. In more stable contexts, poor sleep may distinguish patients at greater risk of poor pain outcomes. In Gaza, sleep problems may be a shared environmental and psychological burden. Clinically, sleep should still be assessed, but its interpretation needs to account for the wider circumstances in which patients are living.

### Sex differences in disability, fatigue and psychological distress

Female participants experienced a greater burden in several domains. In the neck pain group, females reported higher fatigue and psychological distress than males. In the LBP group, females reported higher disability and psychological distress. These findings are partly consistent with previous chronic LBP research, where anxiety and depression were more common among female patients [44].

The findings also align with broader evidence that chronic pain places a disproportionate burden on women [45]. In the current study, this burden was not identical across pain locations. In neck pain, the main sex differences were fatigue and psychological distress. In LBP, the main differences were disability and psychological distress. This suggests that sex-sensitive assessment should consider both pain location and outcome domain.

In the Gaza context, these differences may be intensified by social and environmental pressures. Women may carry increased caregiving responsibilities, household demands and displacement-related burdens, while also facing limited access to support. These pressures may amplify physical strain and psychological distress. Therefore, the sex differences observed in this study should not be reduced to biological explanations alone. They are likely shaped by both sex-related and context-specific factors.

The exploratory interaction analyses suggested that sex may modify the relationship between pain status and both fatigue and psychological distress in the neck pain group. This may indicate that the acute-to-chronic difference in these outcomes is not identical in males and females. However, these analyses were exploratory and the study was not powered primarily for interaction testing. The findings should therefore be treated as hypothesis-generating rather than confirmatory.

### Clinical implications

The findings support early multidimensional assessment of patients presenting with spinal pain. Disability measures remain important, but this study suggests that fatigue and psychological distress should also be assessed routinely, particularly when symptoms persist beyond the acute stage. Fatigue was the most consistent factor associated with chronic pain status across the separate regression models, making it a potentially useful clinical marker.

The findings also support a biopsychosocial approach to spinal pain management. Physical therapy that focuses only on local spinal symptoms may be insufficient for patients with chronic pain, high fatigue or psychological distress. Evidence from multidisciplinary chronic LBP management indicates that anxiety and depression can influence treatment outcomes [46]. Sleep should also remain part of assessment, as poor sleep quality has been associated with chronic neck pain [47].

The PROMIS Sleep Disturbance Short Form has demonstrated reliability and responsiveness in chronic pain research, although not specifically in spinal pain populations [48]. The PHQ-4 also has strong psychometric support as a brief measure of anxiety and depression symptoms in general adult populations [49]. These instruments are practical and can help identify patients who may need additional support, closer follow-up or referral where services are available.

In Gaza and similar settings, these recommendations need to be realistic. Rehabilitation services may be disrupted, and access to specialist mental health support may be limited. Even so, simple screening for disability, fatigue, distress and sleep problems may help clinicians identify patients at higher risk of poor recovery and tailor care more appropriately.

### Strengths and limitations

This study has several strengths. It included a relatively large sample of 410 participants, with balanced acute and chronic subgroups for both neck pain and LBP. It also assessed several clinically relevant domains at the same time, including disability, fatigue, psychological distress and sleep disturbance. The use of established outcome measures strengthens the clinical relevance of the findings. Another important strength is the setting, as evidence from Gaza and similar conflict-affected contexts remains limited.

Several limitations should also be acknowledged. First, the cross-sectional design prevents causal inference. The regression findings should therefore be described as factors associated with chronic pain status, not as proof that these factors predict or cause chronicity. Second, the study was conducted at a single centre, which may limit generalisability. Third, important potential confounders, including pain intensity, BMI, medication use, physical activity, smoking status and trauma exposure, were not systematically controlled. This is particularly important because pain intensity may partly explain the relationships between chronicity, disability, fatigue and psychological distress. Fourth, the NDI and ODI were adapted by excluding one contextually non-applicable item from each scale: the NDI driving item and the ODI travelling item. This was intended to reduce potential measurement error because inability to drive or travel during the conflict could reflect restricted access to vehicles, fuel, safety and ordinary movement rather than pain-related disability. However, because the original instruments are standard ten-item measures [27,28], and adaptation of self-report measures should consider experiential relevance and content validity [50,51], this adaptation may limit direct comparison with studies using the full versions. Finally, the study included several secondary and exploratory analyses. These findings, particularly sex-by-pain-status interactions and sensitivity analyses, should be interpreted cautiously because formal correction for multiple testing was not applied across all comparisons.

The conflict context is both a strength and a limitation. It makes the study highly relevant, but it also introduces factors that are difficult to measure and adjust for. Displacement, disrupted sleep, insecurity, physical overexertion and limited access to rehabilitation may all have influenced the findings. Finally, consecutive sampling may introduce selection bias because the sample included patients who were able to access outpatient physiotherapy during a period of severe healthcare disruption.

### Future research

Future studies should use longitudinal designs to examine whether disability, fatigue, psychological distress and sleep disturbance contribute to the transition from acute to chronic spinal pain. Multi-centre studies would help determine whether the findings are generalisable beyond Nasser Medical Complex. Future research should also include pain intensity, medication use, BMI, physical activity, sleep environment, displacement status and trauma exposure as potential confounders or effect modifiers.

Intervention studies are also needed to test whether early screening and targeted management of fatigue and psychological distress can reduce disability and improve recovery among patients with acute spinal pain. Given the sex differences observed in this study, future research should also examine whether sex-sensitive and context-sensitive rehabilitation approaches improve outcomes for women and men in conflict-affected settings.

## Conclusions

This comparative cross-sectional study found that chronic neck pain and chronic LBP were associated with significantly higher disability, fatigue and psychological distress than acute pain. Fatigue showed the most consistent association with chronic pain status across both spinal regions, while disability was higher in chronic pain groups and remained independently associated with chronicity in the neck pain model. Psychological distress was also higher in chronic pain groups, although it did not remain independently associated with chronic pain status after adjustment in the location-specific regression models. Sleep disturbance did not differ significantly between acute and chronic groups, although it was correlated with fatigue, disability and psychological distress.

These findings support the need for early multidimensional assessment of patients presenting with spinal pain, including routine screening for fatigue, functional limitation and psychological distress. This may be particularly important in conflict-affected and low-resource settings, where displacement, insecurity and limited access to rehabilitation may intensify both physical and emotional burden. However, because the study was cross-sectional, the findings should be interpreted as associations rather than evidence that these factors cause or predict chronicity. Longitudinal and multi-centre studies are needed to clarify temporal relationships and to evaluate whether early biopsychosocial interventions can reduce progression from acute to chronic spinal pain.

## List of abbreviations

BMI: Body mass index
CI: Confidence interval
FSS: Fatigue Severity Scale
ICF: International Classification of Functioning, Disability and Health
LBP: Low back pain
NDI: Neck Disability Index
ODI: Oswestry Disability Index
OR: Odds ratio
PHQ-4: Patient Health Questionnaire-4
PROMIS: Patient-Reported Outcomes Measurement Information System
SD: Standard deviation
SE: Standard error

## Declarations

## Ethics approval and consent to participate

Ethical approval was granted by the Helsinki Committee (HC) for ethical approval, Palestinian Health Research Council (PHRC), Gaza Strip, Palestine (approval number PHRC/HC/1240/25; 04 Aug 2025). Administrative approval was obtained from the Ministry of Health, State of Palestine (Human Resources Development Directorate; correspondence number 25114150; 13 Aug 2025). Permission to conduct the research was also obtained from Al-Azhar University (dated: 31 July 2025). Informed consent was obtained from all participants before participation. De-identified study data were stored securely and were accessible only to the research team.

## Consent for publication

Not applicable.

## Availability of data and materials

The datasets generated and analysed during the current study are not publicly available because public sharing was not included in the ethical approval and because the dataset contains sensitive clinical information from a conflict-affected setting. De-identified data are available from the corresponding author on reasonable request, subject to ethical and institutional approvals.

## Authors’ contributions

MaS contributed to data collection, analysis and interpretation. AAN and MSh contributed to study conception and design and revised the manuscript for intellectual content. RA contributed to statistical analysis, data interpretation and critical revision of clinical aspects of the manuscript. KM provided senior oversight, led manuscript development, contributed to data interpretation and critically revised the manuscript for scientific rigour. All authors contributed to drafting and critical revision of the manuscript. All authors approved the final version of the manuscript and agree to be accountable for all aspects of the work.

## Acknowledgements

For the purpose of open access, the authors have applied a Creative Commons Attribution (CC BY) licence to any Author Accepted Manuscript version arising from this submission. The authors thank Dr Ihab Naser, Dean of the Faculty of Applied Medical Sciences, Al-Azhar University–Gaza, Dr Musab Al-Dabbas, and Ms Josie Rodgers, Royal Devon University Healthcare NHS Foundation Trust, Exeter, for their support. The authors also acknowledge the staff of the outpatient physical therapy department at Nasser Medical Complex, Khan Younis, for facilitating data collection.

## Additional files

Additional file 1. Supplementary materials (.pdf). Supplementary Tables S1-S10 and the study questionnaire.

