## Supplementary materials for "Disability, Fatigue, and Mental Health in Acute versus Chronic Spinal Pain Patients in the Gaza Strip: A Comparative Cross-Sectional Study"

### Supplementary Appendix 1. Supplementary tables

**Supplementary Table S1.** Sociodemographic and clinical characteristics of the study participants

| Characteristic | Neck pain (n=204) | Low back pain (n=206) | Total (N=410) | % of total |
|---|---|---|---|---|
| **Sex** | | | | |
| Male | 64 | 81 | 145 | 35.4% |
| Female | 140 | 125 | 265 | 64.6% |
| **Employment status** | | | | |
| Employed | 95 | 88 | 183 | 44.6% |
| Unemployed | 109 | 118 | 227 | 55.4% |
| **Age group (years)** | | | | |
| 18-20 | 24 | 9 | 33 | 8.0% |
| 21-30 | 78 | 59 | 137 | 33.4% |
| 31-40 | 57 | 60 | 117 | 28.5% |
| 41-50 | 45 | 78 | 123 | 30.0% |
| **Pain duration** | | | | |
| Acute (≤12 weeks) | 101 | 102 | 203 | 49.5% |
| Chronic (>12 weeks) | 103 | 104 | 207 | 50.5% |
| **Previous physiotherapy for current complaint** | | | | |
| No | 171 | 127 | 298 | 72.7% |
| Yes | 33 | 79 | 112 | 27.3% |

**Supplementary Table S2.** Sex differences in disability, fatigue and mental health outcomes in the neck pain group

| Variable | Male Mean ± SD | Female Mean ± SD | t | p-value | Cohen's d |
|---|---|---|---|---|---|
| NDI | 16.11 ± 8.78 (n=64) | 17.16 ± 8.03 (n=140) | -0.846 | 0.399 | 0.13 |
| FSS | 29.98 ± 13.36 (n=64) | 37.17 ± 12.44 (n=139) | -3.734 | <0.001 | 0.56 |
| PHQ-4 | 4.36 ± 3.27 (n=64) | 5.76 ± 3.39 (n=139) | -2.757 | 0.006 | 0.42 |
| PROMIS | 24.52 ± 3.91 (n=64) | 25.49 ± 5.83 (n=138) | -1.219 | 0.224 | 0.18 |

**Supplementary Table S3.** Sex differences in disability, fatigue and mental health outcomes in the low back pain group

| Variable | Male Mean ± SD | Female Mean ± SD | t | p-value | Cohen's d |
|---|---|---|---|---|---|
| ODI | 13.25 ± 6.68 (n=81) | 15.66 ± 7.14 (n=125) | -2.433 | 0.016 | 0.35 |
| FSS | 34.69 ± 14.74 (n=81) | 36.14 ± 13.58 (n=125) | -0.725 | 0.469 | 0.10 |
| PHQ-4 | 4.46 ± 3.56 (n=81) | 5.62 ± 3.66 (n=125) | -2.260 | 0.025 | 0.32 |
| PROMIS | 24.90 ± 6.62 (n=81) | 25.43 ± 5.62 (n=125) | -0.617 | 0.538 | 0.09 |

**Supplementary Table S4.** Pearson correlation coefficients and *p*-values among outcome variables in the neck pain group

| Variable | PROMIS | FSS | NDI | PHQ-4 |
|---|---|---|---|---|
| PROMIS | - | 0.205 (*p*=0.003) | 0.209 (*p*=0.003) | 0.220 (*p*=0.002) |
| FSS | 0.205 (*p*=0.003) | - | 0.577 (*p*<0.001) | 0.657 (*p*<0.001) |
| NDI | 0.209 (*p*=0.003) | 0.577 (*p*<0.001) | - | 0.568 (*p*<0.001) |
| PHQ-4 | 0.220 (*p*=0.002) | 0.657 (*p*<0.001) | 0.568 (*p*<0.001) | - |

**Note:** Values are Pearson correlation coefficients with *p*-values in parentheses. Pairwise n ranged from 201 to 203 because analyses were based on available valid outcome data. PROMIS: PROMIS Sleep Disturbance Short Form 8a raw summed score; FSS: Fatigue Severity Scale; NDI: Neck Disability Index; PHQ-4: Patient Health Questionnaire-4.

**Supplementary Table S5.** Pearson correlation coefficients and *p*-values among outcome variables in the low back pain group

| Variable | PROMIS | FSS | ODI | PHQ-4 |
|---|---|---|---|---|
| PROMIS | - | 0.288 ($p$<0.001) | 0.375 ($p$<0.001) | 0.369 ($p$<0.001) |
| FSS | 0.288 ($p$<0.001) | - | 0.604 ($p$<0.001) | 0.553 ($p$<0.001) |
| ODI | 0.375 ($p$<0.001) | 0.604 ($p$<0.001) | - | 0.481 ($p$<0.001) |
| PHQ-4 | 0.369 ($p$<0.001) | 0.553 ($p$<0.001) | 0.481 ($p$<0.001) | - |

**Note:** Values are Pearson correlation coefficients with *p*-values in parentheses. n=206 for all pairwise correlations. PROMIS: PROMIS Sleep Disturbance Short Form 8a raw summed score; FSS: Fatigue Severity Scale; ODI: Oswestry Disability Index; PHQ-4: Patient Health Questionnaire-4.

**Supplementary Table S6.** Adjusted linear regression models comparing chronic versus acute pain status

Positive coefficients indicate higher scores in chronic pain groups. Models were adjusted for sex, age group, employment status and previous physiotherapy.

| Location | Outcome | n | Adjusted B for chronic pain | 95% CI | p-value | Model $R^2$ |
|---|---|---|---|---|---|---|
| Neck pain | NDI | 204 | 8.84 | 6.70 to 10.98 | <0.001 | 0.353 |
| Neck pain | FSS | 203 | 13.11 | 9.84 to 16.38 | <0.001 | 0.403 |
| Neck pain | PHQ-4 | 203 | 3.27 | 2.33 to 4.21 | <0.001 | 0.272 |
| Neck pain | PROMIS | 202 | -0.57 | -2.23 to 1.10 | 0.502 | 0.055 |
| Low back pain | ODI | 206 | 3.33 | 1.52 to 5.14 | <0.001 | 0.273 |
| Low back pain | FSS | 206 | 10.49 | 6.95 to 14.03 | <0.001 | 0.300 |
| Low back pain | PHQ-4 | 206 | 1.84 | 0.84 to 2.83 | <0.001 | 0.189 |
| Low back pain | PROMIS | 206 | 0.39 | -1.35 to 2.12 | 0.661 | 0.090 |

**Supplementary Table S7.** Clinical-threshold analysis for fatigue and psychological distress by pain duration

**S7a.** Clinically significant fatigue by pain duration

| Location | Pain status | Below threshold FSS <36 | Clinically significant fatigue FSS ≥36 | p-value |
|---|---|---|---|---|
| Neck pain | Acute | 76 | 25 | <0.001 |
| Neck pain | Chronic | 17 | 85 | |
| Low back pain | Acute | 71 | 31 | <0.001 |
| Low back pain | Chronic | 35 | 69 | |

**Note**: FSS ≥36 corresponds to a mean item score of ≥4, indicating clinically significant fatigue.

**S7b.** PHQ-4 psychological distress categories by pain duration

| Location | Pain status | Normal 0-2 | Mild 3-5 | Moderate 6-8 | Severe 9-12 | p-value |
|---|---|---|---|---|---|---|
| Neck pain | Acute | 53 | 25 | 10 | 12 | <0.001 |
| Neck pain | Chronic | 8 | 23 | 41 | 31 | |
| Low back pain | Acute | 49 | 28 | 10 | 15 | <0.001 |
| Low back pain | Chronic | 20 | 29 | 23 | 32 | |

**Supplementary Table S8.** Non-parametric sensitivity analyses comparing acute and chronic pain groups

| Location | Outcome | Acute n | Chronic n | Mann-Whitney U | p-value |
|---|---|---|---|---|---|
| Neck pain | NDI | 101 | 103 | 1677.0 | <0.001 |
| Neck pain | FSS | 101 | 102 | 2002.5 | <0.001 |
| Neck pain | PHQ-4 | 100 | 103 | 2058.5 | <0.001 |
| Neck pain | PROMIS | 100 | 102 | 5255.5 | 0.708 |
| Low back pain | ODI | 102 | 104 | 2935.5 | <0.001 |
| Low back pain | FSS | 102 | 104 | 2497.0 | <0.001 |
| Low back pain | PHQ-4 | 102 | 104 | 3226.5 | <0.001 |
| Low back pain | PROMIS | 102 | 104 | 4374.5 | 0.029 |

Note: Mann-Whitney U tests were used as non-parametric sensitivity analyses. PROMIS, PROMIS Sleep Disturbance Short Form 8a raw summed score; FSS, Fatigue Severity Scale; PHQ-4, Patient Health Questionnaire-4; NDI, Neck Disability Index; ODI, Oswestry Disability Index.

**Supplementary Table S9**. Spearman rank correlations among outcome variables

**S9a.** Neck pain group

| Variables | n | Spearman's rho | p-value |
|---|---|---|---|
| PROMIS and FSS | 202 | 0.141 | 0.046 |
| PROMIS and NDI | 202 | 0.136 | 0.053 |
| PROMIS and PHQ-4 | 201 | 0.130 | 0.067 |
| FSS and NDI | 203 | 0.562 | <0.001 |
| FSS and PHQ-4 | 202 | 0.657 | <0.001 |
| NDI and PHQ-4 | 203 | 0.614 | <0.001 |

**S9b.** Low back pain group

| Variables | n | Spearman's rho | p-value |
|---|---|---|---|
| PROMIS and FSS | 206 | 0.323 | <0.001 |
| PROMIS and ODI | 206 | 0.392 | <0.001 |
| PROMIS and PHQ-4 | 206 | 0.356 | <0.001 |
| FSS and ODI | 206 | 0.647 | <0.001 |
| FSS and PHQ-4 | 206 | 0.573 | <0.001 |
| ODI and PHQ-4 | 206 | 0.503 | <0.001 |

**Supplementary Table S10.** Exploratory sex-by-pain-status interaction models

The interaction term tests whether the difference between acute and chronic pain differs by sex. Models were adjusted for age group, employment status and previous physiotherapy.

| Location | Outcome | n | Interaction B | 95% CI | p-value |
|---|---|---|---|---|---|
| Neck pain | NDI | 204 | -3.37 | -7.56 to 0.82 | 0.115 |
| Neck pain | FSS | 203 | -6.76 | -13.14 to -0.38 | 0.038 |
| Neck pain | PHQ-4 | 203 | -2.36 | -4.18 to -0.54 | 0.011 |
| Neck pain | PROMIS | 202 | 1.89 | -1.38 to 5.17 | 0.255 |
| Low back pain | ODI | 206 | -1.15 | -4.67 to 2.36 | 0.519 |
| Low back pain | FSS | 206 | -3.88 | -10.74 to 2.97 | 0.265 |
| Low back pain | PHQ-4 | 206 | -1.05 | -2.97 to 0.88 | 0.285 |

| | | | | | |
|---|---|---|---|---|---|
| Low back pain | PROMIS | 206 | 2.41 | -0.94 to 5.76 | 0.157 |

### Supplementary Appendix 2. Study questionnaire

### Questionnaire for Sociodemographic Data

| Sociodemographic Data | |
|---|---|
| Sex | ☐Male ☐ Female |
| Employment Status: | ☐Work ☐ Do not Work. |
| Age | ☐18-20 ☐ 21-30 ☐31-40 ☐41-50 |
| Pain duration: | ☐≤12weeks ☐ >12 weeks |
| Did you have a previous physical therapy session for your problem? | ☐ Yes ☐ No |

#### Modified Oswestry LBP Disability Index (ODI)

**Pain Intensity**

_____The pain is mild and comes and goes.
_____The pain is mild and does not vary much.
_____The pain is moderate and comes and goes.
_____The pain is moderate and does not vary much.
_____The pain is severe and comes and goes.
_____The pain is severe and does not vary much.

**Personal Care (Washing, Dressing, etc.)**

_____I do not have to change the way I wash and dress myself to avoid pain.
_____I do not normally change the way I wash or dress myself, even though it causes some pain.
_____Washing and dressing increase my pain, but I can do it without changing my way of doing it.
_____Washing and dressing increases my pain, and I find it necessary to change the way I do it.
_____Because of my pain I am partially unable to wash and dress without help.
_____Because of my pain I am completely unable to wash or dress without help.

**Lifting**

_____I can lift heavy weights without increased pain.
_____I can lift heavy weights but it causes increased pain
_____Pain prevents me from lifting heavy weights off of the floor, but I can manage if they are conveniently positioned (ex. on a table, etc.).
_____Pain prevents me from lifting heavy weights off of the floor, but I can manage light to medium weights if they are conveniently positioned.
_____I can lift only very light weights.
_____I can not lift or carry anything at all.

**Walking**

_____I have no pain when walking.
_____I have pain when walking, but I can still walk my required normal distances.
_____Pain prevents me from walking long distances.
_____Pain prevents me from walking intermediate distances.
_____Pain prevents me from walking even short distances.
_____Pain prevents me from walking at all.

**Sitting**

_____Sitting does not cause me any pain.
_____I can only sit as long as I like providing that I have my choice of seating surfaces.
_____Pain prevents me from sitting for more than 1 hour.

| |
|---|
| _____Pain prevents me from sitting for more than 1/2 hour.<br>_____Pain prevents me from sitting for more than 10 minutes.<br>_____Pain prevents me from sitting at all. |
| **Standing**<br>_____I can stand as long as I want without increased pain.<br>_____I can stand as long as I want, but my pain increases with time.<br>_____Pain prevents me from standing for more than 1 hour.<br>_____Pain prevents me from standing for more than 1/2 hour.<br>_____Pain prevents me from standing for more than 10 minutes.<br>_____I avoid standing because it increases my pain right away.<br><br>**Sleeping**<br>_____I get no pain when I am in bed.<br>_____I get pain in bed, but it does not prevent me from sleeping well.<br>_____Because of my pain, my sleep is only 3/4 of my normal amount.<br>_____Because of my pain, my sleep is only 1/2 of my normal amount.<br>_____Because of my pain, my sleep is only 1/4 of my normal amount.<br>_____Pain prevents me from sleeping at all.<br><br>**Social Life**<br>_____My social life is normal and does not increase my pain.<br>_____My social life is normal, but it increases my level of pain.<br>_____Pain prevents me from participating in more energetic activities (ex., sports, dancing, etc.)<br>_____Pain prevents me from going out very often.<br>_____Pain has restricted my social life to my home.<br>_____I have hardly any social life because of my pain.<br><br>**Traveling**<br>_____I get no increased pain when traveling.<br>_____I get some pain while traveling, but none of my usual forms of travel make it any worse.<br>_____I get increased pain while traveling, but it does not cause me to seek alternative forms of travel.<br>_____I get increased pain while traveling which causes me to seek alternative forms of travel.<br>_____My pain restricts all forms of travel except that which is done while I am lying down.<br>_____My pain restricts all forms of travel.<br><br>**Employment/Homemaking**<br>_____My normal job/homemaking activities do not cause pain.<br>_____My normal job/homemaking activities increase my pain, but I can still perform all that is required of me.<br>_____I can perform most of my job/homemaking duties, but pain prevents me from performing more physically stressful activities (ex. lifting, vacuuming)<br>_____Pain prevents me from doing anything but light duties.<br>_____Pain prevents me from doing even light duties.<br>_____Pain prevents me from performing any job or homemaking chores. |
| **Total score: _____ /45** |

Scoring note: The travelling item was included in the administered questionnaire but was excluded from the ODI total score because it was not consistently applicable during the conflict context in Gaza. The ODI total score was therefore calculated from nine scored items, with a possible raw score range of 0-45. Higher scores indicate greater disability.

### Neck Disability Index

| **Section 1: Pain Intensity** |
|---|
| ☐ I have no pain at the moment. |
| ☐ The pain is very mild at the moment. |
| ☐ The pain is moderate at the moment. |

| |
|---|
| ☐ The pain is fairly severe at the moment. |
| ☐ The pain is very severe at the moment. |
| ☐ The pain is the worst imaginable at the moment. |
| **Section 2: Personal Care (Washing, Dressing, etc.)** |
| ☐ I can look after myself normally without causing extra pain. |
| ☐ I can look after myself normally, but it causes extra pain. |
| ☐ It is painful to look after myself, and I am slow and careful. |
| ☐ I need some help, but I can manage most of my personal care. |
| ☐ I need help every day in most aspects of self-care. |
| ☐ I do not get dressed; I wash with difficulty and stay in bed. |
| **Section 3: Lifting** |
| ☐ I can lift heavy weights without extra pain. |
| ☐ I can lift heavy weights, but it gives extra pain |
| ☐ Pain prevents me lifting heavy weights off the floor, but I can manage if they are conveniently placed, for example on a table. |
| ☐ Pain prevents me from lifting heavy weights, but I can manage light to medium weights if they are conveniently positioned. |
| ☐ I can only lift very light weights. |
| ☐ I cannot lift or carry anything. |
| **Section 4: Reading** |
| ☐ I can read as much as I want to with no pain in my neck. |
| ☐ I can read as much as I want to with slight pain in my neck. |
| ☐ I can read as much as I want with moderate pain in my neck. |
| ☐ I can't read as much as I want because of moderate pain in my neck. |
| ☐ I can hardly read at all because of severe pain in my neck. |
| ☐ I cannot read at all. |
| **Section 5: Headaches** |
| ☐ I have no headaches at all. |
| ☐ I have slight headaches, which come infrequently. |
| ☐ I have moderate headaches, which come infrequently. |
| ☐ I have moderate headaches, which come frequently. |
| ☐ I have severe headaches, which come frequently. |
| ☐ I have headaches almost all the time. |
| **Section 6: Concentration** |

| |
|---|
| ☐ I can concentrate fully when I want to with no difficulty. |
| ☐ I can concentrate fully when I want to, with slight difficulty. |
| ☐ I have a fair degree of difficulty in concentrating when I want to. |
| ☐ I have a lot of difficulty in concentrating when I want to. |
| ☐ I have a great deal of difficulty concentrating when I want to. |
| ☐ I cannot concentrate at all. |
| **Section 7: Work** |
| ☐ I can do as much work as I want to. |
| ☐ I can only do my usual work, but no more. |
| ☐ I can do most of my usual work, but no more |
| ☐ I cannot do my usual work. |
| ☐ I can hardly do any work at all. |
| ☐ I can’t do any work at all. |
| **Section 8: Driving** |
| ☐ I can drive my car without any neck pain. |
| ☐ I can drive my car as long as I want with slight pain in my neck. |
| ☐ I can drive my car as long as I want with moderate pain in my neck. |
| ☐ I can’t drive my car as long as I want because of moderate pain in my neck. |
| ☐ I can hardly drive at all because of severe pain in my neck. |
| ☐ I can’t drive my car at all. |
| **Section 9: Sleeping** |
| ☐ I have no trouble sleeping. |
| ☐ My sleep is slightly disturbed (less than 1 hr sleepless). |
| ☐ My sleep is mildly disturbed (1-2 hrs sleepless). |
| ☐ My sleep is moderately disturbed (2-3 hrs sleepless). |
| ☐ My sleep is greatly disturbed (3-5 hrs sleepless). |
| ☐ My sleep is completely disturbed (5-7 hrs sleepless). |
| **Section 10: Recreation** |
| ☐ I am able to engage in all my recreation activities with no neck pain at all. |
| ☐ I am able to engage in all my recreation activities, with some pain in my neck. |
| ☐ I am able to engage in most, but not all of my usual recreation activities because of pain in my neck. |
| ☐ I am able to engage in a few of my usual recreation activities because of pain in my neck. |
| ☐ I can hardly do any recreation activities because of pain in my neck. |

| |
|---|
| ☐ I can't do any recreation activities at all. |
| **Score: —/45** |

Scoring note: The driving item was included in the administered questionnaire but was excluded from the NDI total score because it was not consistently applicable during the conflict context in Gaza. The NDI total score was therefore calculated from nine scored items, with a possible raw score range of 0-45. Higher scores indicate greater disability.

### Fatigue Severity Scale

| **During the past week, I have found that:** | **Strongly Disagree** | | | **Neither Agree Nor disagree.** | | | **Strongly Agree** |
|---|---|---|---|---|---|---|---|
| 1. My motivation is lower when I am fatigued. | 1 | 2 | 3 | 4 | 5 | 6 | 7 |
| 2. Exercise brings on my fatigue. | 1 | 2 | 3 | 4 | 5 | 6 | 7 |
| 3. I am easily fatigued. | 1 | 2 | 3 | 4 | 5 | 6 | 7 |
| 4. Fatigue interferes with physical functioning. | 1 | 2 | 3 | 4 | 5 | 6 | 7 |
| 5. Fatigue causes frequent problems for me. | 1 | 2 | 3 | 4 | 5 | 6 | 7 |
| 6. My fatigue prevents sustained physical functioning. | 1 | 2 | 3 | 4 | 5 | 6 | 7 |
| 7. Fatigue interferes with carrying out specific duties and responsibilities. | 1 | 2 | 3 | 4 | 5 | 6 | 7 |
| 8. Fatigue is among my three most disabling symptoms. | 1 | 2 | 3 | 4 | 5 | 6 | 7 |
| 9. Fatigue interferes with my work, family or social life. | 1 | 2 | 3 | 4 | 5 | 6 | 7 |
| **Total Score:** | | | | | | | |

### PHQ-4: The Four-Item Patient Health

### Questionnaire For Anxiety and Depression

| Over the last two weeks, how often have you been bothered by the following problems? | Not at all | Several days | More than half the days | Nearly every day |
|---|---|---|---|---|
| Feeling nervous, anxious or on edge | 0 | 1 | 2 | 3 |
| Not being able to stop or control worrying | 0 | 1 | 2 | 3 |

| | | | | |
|---|---|---|---|---|
| Feeling down, depressed or hopeless | 0 | 1 | 2 | 3 |
| Little interest or pleasure in doing things | 0 | 1 | 2 | 3 |
| TOTAL SCORE | | | | |

### PROMIS-Sleep Disturbance-Short Form

| | | | | | | **Clinician use** |
|---|---|---|---|---|---|---|
| **In the past SEVEN (7) DAYS….** | | | | | | |
| | **Not at all** | **A little bit** | **Somewhat** | **Quite a bit** | **Very much** | |
| 1. My sleep was restless | ☐ 1 | ☐ 2 | ☐ 3 | ☐ 4 | ☐ 5 | |
| 2. I was satisfied with my sleep | ☐ 5 | ☐ 4 | ☐ 3 | ☐ 2 | ☐ 1 | |
| 3. My sleep was refreshing | ☐ 5 | ☐ 4 | ☐ 3 | ☐ 2 | ☐ 1 | |
| 4. I had difficulty falling asleep | ☐ 1 | ☐ 2 | ☐ 3 | ☐ 4 | ☐ 5 | |
| **In the past SEVEN (7) DAYS….** | | | | | | |
| | **Never** | **Rarely** | **Sometimes** | **Often** | **Always** | |
| 5. I had trouble staying asleep | ☐ 1 | ☐ 2 | ☐ 3 | ☐ 4 | ☐ 5 | |
| 6. I had trouble sleeping | ☐ 1 | ☐ 2 | ☐ 3 | ☐ 4 | ☐ 5 | |
| 7. I got enough sleep | ☐ 5 | ☐ 4 | ☐ 3 | ☐ 2 | ☐ 1 | |
| **In the past SEVEN (7) DAYS….** | | | | | | |
| | **Very poor** | **Poor** | **Fair** | **Good** | **Very good** | |
| 8. My sleep quality was… | ☐ 5 | ☐ 4 | ☐ 3 | ☐ 2 | ☐ 1 | |
| | | | | | **Total Score:** | |
